# The Socioeconomic Gradient in Epigenetic Aging Clocks: Evidence from the Multi-Ethnic Study of Atherosclerosis and the Health and Retirement Study

**DOI:** 10.1101/2021.03.01.21252660

**Authors:** Lauren L. Schmitz, Wei Zhao, Scott M. Ratliff, Julia Goodwin, Jiacheng Miao, Qiongshi Lu, Xiuqing Guo, Kent D. Taylor, Jingzhong Ding, Yongmei Liu, Morgan Levine, Jennifer A. Smith

## Abstract

Epigenetic clocks have been widely used to predict disease risk in multiple tissues or cells. Their success as a measure of biological aging has prompted research on the connection between epigenetic pathways of aging and the socioeconomic gradient in health and mortality. However, studies examining social correlates of epigenetic aging have yielded inconsistent results. We conducted a comprehensive, comparative analysis of associations between various dimensions of socioeconomic status (SES) (education, income, wealth, occupation, neighborhood environment, and childhood SES) and eight epigenetic clocks in two large U.S. aging studies: The Multi-Ethnic Study of Atherosclerosis (MESA) (n=1,211) and the Health and Retirement Study (HRS) (n=4,018). In both studies, we found robust associations between SES measures in adulthood and the GrimAge and DunedinPoAm clocks (Bonferroni corrected *p*-value<0.01). In the HRS, significant associations with the Levine and Yang clocks are also evident. These associations are only partially mediated by smoking, alcohol consumption, and obesity, which suggests that differences in health behaviors alone cannot explain the SES gradient in epigenetic aging. Further analyses revealed concurrent associations between polygenic risk for accelerated intrinsic epigenetic aging, SES, and the Levine clock, indicating that genetic predisposition and social disadvantage may contribute independently to faster epigenetic aging.

## Introduction

Accumulating evidence suggests that health inequalities at older ages are triggered by socioeconomic stressors and amplified across the life course through biological and behavioral channels.^1–4^ However, the pathways through which socioeconomic status (SES) becomes biologically embedded and affects health are not well understood, in part because social adversity is linked to a broad range of conditions and diseases that affect multiple physiological systems and tissue types.^5^ In addition, the lack of epidemiological studies with biological and socioeconomic measures across the life course has made it challenging to separate biological correlates of social adversity from its consequences.

In recent years, the expansion of epigenomic profiling has enabled further research on dynamic changes in DNA methylation (DNAm) that accompany the aging process in larger population studies. Principal among these advances has been the development of epigenetic clocks as molecular indicators of biological aging.^6–18^ Epigenetic clocks are multivariate weighted sums of cytosine methylation at cytosine-phosphate-guanine (CpG) sites across the genome developed via machine learning to predict chronological age or other phenotypic hallmarks of aging. Epigenetic clock measures accurately predict chronological age,^6, 11–14, 16, 18–20^ and an increasing number of studies have linked deviations between DNAm age and chronological age (i.e, epigenetic age acceleration or EAA) with age-related diseases and mortality.^10, 21–38^ These findings suggest that epigenetic clocks may serve as molecular biomarkers of aging that can be applied to multiple sources of DNA (cells, tissues, and organs).^39^ Consequently, examining differences in EAA across individuals could help determine the mechanisms, processes, and pathways that connect social and biological forces.

Several studies have examined associations between EAA and measures of SES across the life course, including education, occupational position, income, and childhood SES.^26, 40–51^ The vast majority of these studies have utilized the Horvath and Hannum clocks, often referred to as “first generation clocks”. These studies have found inconsistent evidence on the association between socioeconomic attainment and EAA, with some studies finding a negative and significant association with the Hannum clock^42^ and null associations with the Horvath clock.^42, 52, 53^ The most frequently reported null findings were with regards to education and the Horvath clock (see Ryan et al., 2020 for a review). Studies investigating childhood SES have shown more consistent findings: early life socioeconomic disadvantage is associated with EAA in both the Horvath and Hannum clocks.^41, 49, 55, 56^ However, several studies have found a null relationship between childhood SES and EAA as measured by first generation clocks.^53, 57–59^ More recently, studies have begun to explore SES associations with second generation clocks trained on phenotypic biomarkers of aging. These studies have found more consistent associations between socioeconomic position and the Levine, GrimAge, and DunedinPoAm clocks,^9, 10, 48, 59–62^ suggesting that these clocks may be capturing pathways of aging that are more proximal to social conditions. Overall, the lack of consistent evidence for most SES exposures is likely driven by differences in study design, cohort norms, underlying sample characteristics, lack of statistical power, and positive publication bias.^54^ In addition, few studies have examined correlations between SES and second-generation clocks, with only one study to date looking at the correlation between education and all 13 clocks in adults.^62^ Many studies also do not account for the potential mediating relationship of lifestyle factors (namely smoking, body mass index (BMI), and alcohol consumption), despite their high correlation with both socioeconomic position and epigenetic profiles.^42, 48, 51, 63–65^ The inconsistency across studies of older adults in particular necessitates further examination of the relationship between epigenetic aging and adult socioeconomic position in multiple, well-powered studies. Moreover, evidence regarding the particular aspects or dimensions of SES that relate to epigenetic clock measures may help future researchers pinpoint epigenetic pathways of aging that are related to the social environment.

In this study, we conducted a comprehensive, comparative analysis of associations between various dimensions of SES and eight epigenetic clocks in two large U.S. aging studies: The Multi-Ethnic Study of Atherosclerosis (MESA) (n=1,211) and the Health and Retirement Study (HRS) (n=4,018). In subsequent analyses, we examined the degree to which these associations are mediated by smoking, BMI, and alcohol consumption. Given that epigenetic aging is to an extent regulated by genetics, we also incorporated polygenic scores (PGS) for intrinsic and extrinsic epigenetic age acceleration (IEAA and EEAA) using findings from a recent genome-wide association study (GWAS).^66^ To our knowledge, this is the first study to use a PGS to account for genetic heterogeneity in EAA in the SES-clock literature.

## Results

### Epigenetic clock characteristics

To date, more than a dozen epigenetic clocks have been developed to quantify biological aging. Here, we give a brief overview of the basic methodology and theoretical interpretation behind the clock measures used in this study (in-depth reviews of the various epigenetic clocks can be found elsewhere).^39, 67^ Importantly, recent research has found that the underlying CpG sites and weights used to construct the clocks display minimal overlap and capture distinct phenomena of epigenetic aging, perhaps because of the different outcome measures, tissues, and populations that were used to develop clock algorithms.^68^ As a result, we tested SES relationships across eight epigenetic clocks that by definition differ in their age correlations in different tissues and cells.

**Table 1** lists the eight clocks examined in this study in their approximate order of publication with information on the training phenotype, tissue type, and number of CpG sites used to derive clock measures. First generation clocks, including Horvath’s original pan-tissue clock and subsequent skin and blood clock (referred to herein as Horvath 1 and Horvath 2), and the Hannum and Lin clocks, used penalized regression methods (i.e., elastic net) to train an epigenetic predictor of chronological age in whole blood or across various tissue and cell types. The epiTOC or Yang clock was developed to approximate a “mitotic clock” that defines the average level of methylation across 385 CpG sites associated with age-related increases in cell division or cell-replication errors.^16^ Subsequent second-generation clocks, including Levine DNAm PhenoAge and Lu’s DNAm GrimAge, incorporated more complex training phenotypes based on both clinical, phenotypic aging measures and mortality risk to more closely capture underlying biological features of accelerated epigenetic aging.^8, 9^ Additionally, unlike previous clocks which are based solely on information from CpG methylation, DNAm GrimAge also includes observed chronological age and self-reported sex as additional variables in the model. More recently, Belsky et al. (2020) developed a clock in the Dunedin Study that used multiple waves of biomarker data on the same cohort between the ages of 26 and 38 to quantify the pace of aging among individuals born in the same year. The resulting DunedinPoAm clock (Dunedin(P)ace(o)f(A)ging(m)ethylation clock) is theorized to capture within individual variation in rate of aging over time, whereas prior clocks were trained to capture cross-sectional differences in biological aging across similarly aged individuals or individuals from different age groups. As such, the DunedinPoAm clock does not strongly track with chronological age.

**Table 1.**
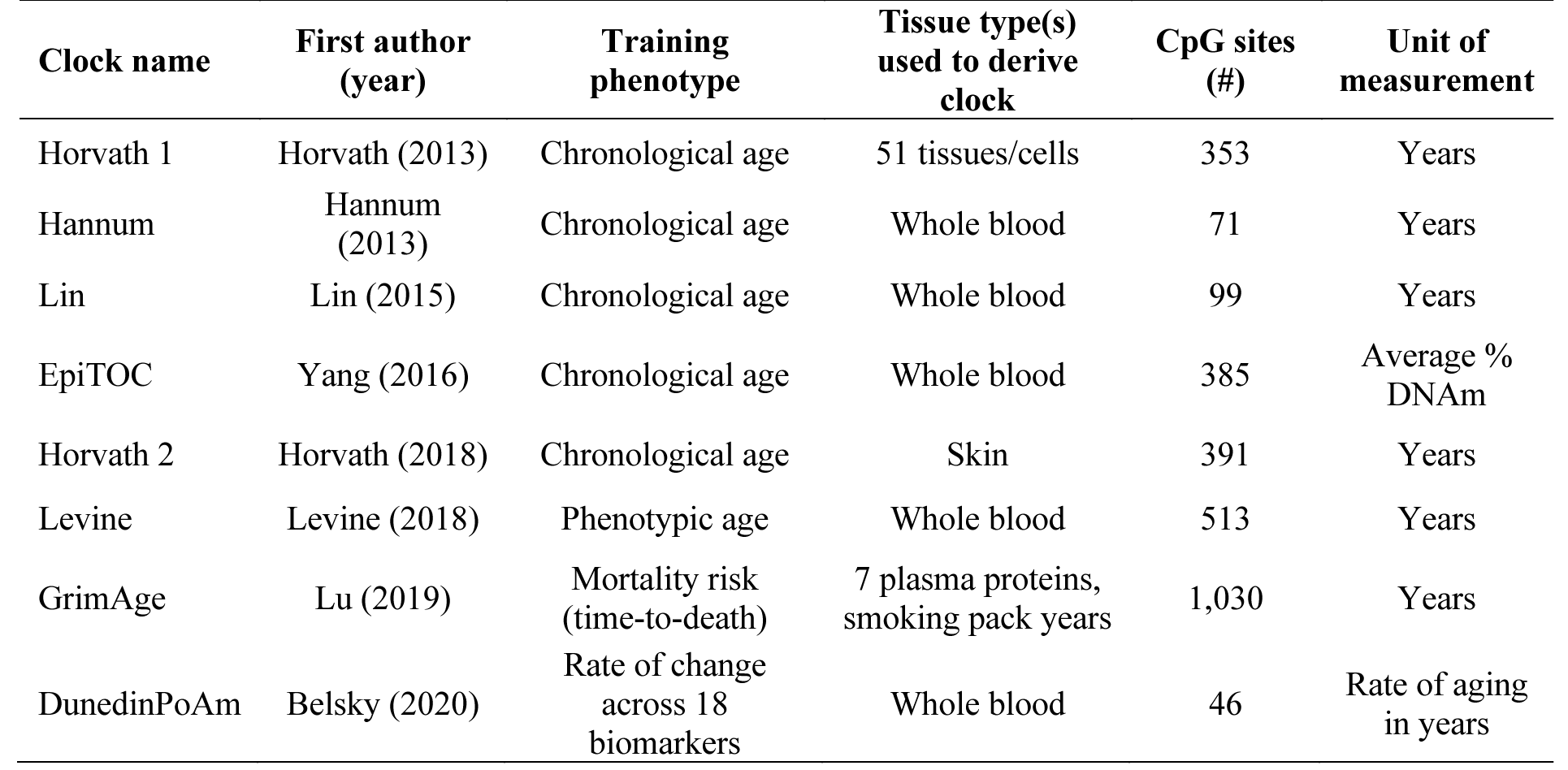
Characteristics of epigenetic aging clocks examined in MESA and HRS

| **Clock name** | **First author**<br>**(year)** | **Training**<br>**phenotype** | **Tissue type(s)**<br>**used to derive**<br>**clock** | **CpG sites**<br>**(#)** | **Unit of**<br>**measurement** |
| --- | --- | --- | --- | --- | --- |
| Horvath 1 | Horvath (2013) | Chronological age | 51 tissues/cells | 353 | Years |
| Hannum | Hannum<br>(2013) | Chronological age | Whole blood | 71 | Years |
| Lin | Lin (2015) | Chronological age | Whole blood | 99 | Years |
| EpiTOC | Yang (2016) | Chronological age | Whole blood | 385 | Average %<br>DNAm |
| Horvath 2 | Horvath (2018) | Chronological age | Skin | 391 | Years |
| Levine | Levine (2018) | Phenotypic age | Whole blood | 513 | Years |
| GrimAge | Lu (2019) | Mortality risk<br>(time-to-death) | 7 plasma proteins,<br>smoking pack years | 1,030 | Years |
| DunedinPoAm | Belsky (2020) | Rate of change<br>across 18<br>biomarkers | Whole blood | 46 | Rate of aging<br>in years |

**Tables S1-S4** in the Supplementary Appendix reflect the overall lack of correlation across clocks and their age-adjusted EAA measures in MESA and the HRS. With the exception of DunedinPoAm, which was not trained on chronological age, and the Yang clock, which was restricted to CpG sites that map to Polycomb group targets (PCGTs) in embryonic stem cells, all other clocks are strongly correlated with chronological age in both samples (*r*=0.71-0.894, *p*<0.0001 for all). After adjusting for age, correlations between the clocks fall significantly; on the high-end EAA correlations range between approximately 0.50 and 0.60 for the Horvath EAA and Hannum EAA measures (*p*<0.0001). On the low end, clock correlations fall to 0.035 (*p*=0.027) or are even negatively correlated (*r*=-0.223, *p*<0.0001) for DunedinPoAm EAA correlations with the Horvath 2 and Yang clocks, respectively.

### Sample characteristics

**Table 2** describes the characteristics of the MESA and HRS samples. Descriptive statistics for time-varying characteristics are reported for the exam or wave that blood was collected with the exception of neighborhood and occupation measures, which represent the average exposure across all available exams or waves. In MESA, wealth was reported at Exam 3. The mean age at blood collection in both samples was approximately 69 years old (MESA: 69.6, SD=9.31; HRS: 69.48, SD=9.63). In MESA, nearly half (50.5%) of participants were female compared to 58% in the HRS. MESA participants were more racially and ethnically diverse compared to HRS participants: 48% of MESA participants report being non-Hispanic White compared to 66% of HRS participants. Overall, sample averages of epigenetic clock measures in MESA and HRS are either strikingly similar or do not differ significantly from one another, perhaps due to the similarities in the age distribution and the average age at blood collection. Average epigenetic age varies between 54.6 and 82.5 depending on the clock algorithm (for clocks that approximate biological age). On average, the Yang epiTOC clock, which estimates average DNAm due to putative cell-replication errors, shows a 7% (SD=0.02) and 9% (SD=0.01) increase in DNAm at these sites in HRS and MESA, respectively. The DunedinPoAm clock, which measures rate of aging in years, approximates that individuals in MESA and the HRS are on average aging 6% (1.06, SD=0.08) and 8% (1.08, SD=0.09) faster biologically compared to their chronological age.

**Table 2.** Descriptive characteristics of study participants by cohort, MESA and HRS

|  | **Mean (SD) or n (%)** |  |
| --- | --- | --- |
|  | **MESA**<br>**(n=1,211)** | **HRS**<br>**(n=4,018)** |
| Age | 69.61 (9.31) | 69.48 (9.63) |
| Female | 612 (50.54) | 2330 (58) |
| Race |  |  |
| Non-Hispanic White | 582 (48.06) | 2,669 (66.46) |
| Non-Hispanic Black | 243 (20.07) | 658 (16.38) |
| Non-Hispanic “other” race | . | 122 (3.04) |
| Hispanic | 386 (31.87) | 567 (14.11) |
| Epigenetic aging clocks |  |  |
| Horvath 1 | 63.86 (8.48) | 65.72 (9.62) |
| Horvath 2 | 69.19 (7.55) | 69.61 (8.85) |
| Hannum | 73.02 (8.55) | 54.59 (9.23) |
| Levine PhenoAge | 71.77 (9.28) | 57.48 (10.08) |
| Lin | 82.54 (10.58) | 58.42 (11.08) |
| Yang epiTOC | 0.09 (0.01) | 0.07 (0.02) |
| GrimAge | 79.40 (8.10) | 68.13 (8.63) |
| DunedinPoAm | 1.06 (0.08) | 1.08 (0.09) |
| SES Index | 2.82 (0.91) | 3.18 (0.93) |
| Occupational disadvantage score | -0.01 (1.01) | 0.02 (1.02) |
| Neighborhood socioeconomic disadvantage score | 0.01 (0.99) | 0.02 (1.01) |
| Neighborhood social environment disadvantage score | -0.03 (0.99) | 0.04 (1.05) |
| Income quintiles |  |  |
| <\$5,000-\$19,999 | 210 (17.34) | 1080 (26.88) |
| \$20,000-\$34,999 | 238 (19.65) | 822 (20.46) |
| \$35,000-\$49,999 | 226 (18.66) | 565 (14.06) |
| \$50,000-\$74,999 | 220 (18.17) | 578 (14.39) |
| \$75,000+ | 275 (22.71) | 973 (24.22) |
| Wealth |  |  |
| Assets in home/land, car, and financial investments | 679 (56.77) | 575 (14.31) |
| Assets in home/land and car or investments | 181 (15.13) | 476 (11.85) |
| Assets in home/land only or car and investments | 88 (7.36) | 1255 (31.23) |
| Assets in financial investments or car and not home/land | 134 (11.2) | 1119 (27.85) |
| No assets | 114 (9.53) | 593 (14.76) |
| Educational attainment |  |  |
| No degree | 175 (14.47) | 675 (16.80) |
| HS degree/GED | 234 (19.35) | 2116 (52.66) |
| Some college | 399 (33) | 259 (6.45) |
| College (BA) | 188 (15.55) | 608 (15.13) |
| Advanced degree | 213 (17.62) | 360 (8.96) |
| Married |  |  |
| Yes | 695 (57.39) | 2511 (62.49) |
| No | 491 (40.55) | 1504 (37.43) |
| Smoking Status |  |  |
| Current | 121 (9.99) | 455 (11.32) |
| Former (>1 year) | 493 (40.71) | 1776 (44.20) |
| Never | 577 (47.65) | 1764 (43.90) |
| Obesity |  |  |
| Normal: BMI≤25 | 246 (20.31) | 1103 (27.45) |
| Overweight: 25<BMI<30 | 456 (37.65) | 1426 (35.49) |
| Obese: BMI≥30 | 508 (41.95) | 1446 (35.99) |
| Drinking |  |  |
| Non-drinker | 786 (64.91) | 1783 (44.38) |
| ≤2 drinks/day | 355 (29.31) | 1844 (45.89) |
| >2 drinks/day | 65 (5.37) | 380 (9.46) |
| Mother: years of education | 10.82 (3.68) | 9.95 (3.91) |
| Father: years of education | 10.74 (3.93) | 9.76 (4.12) |

**Note:** SD=standard deviation. Categories for indicator variables may not sum to 100% if information was missing for some individuals. In the HRS, the non-Hispanic other race category includes individuals who identify as American Indian, Alaskan Native, Asian, or Pacific Islander.

Both samples are socioeconomically diverse. Approximately 15% of MESA respondents have no degree (HRS: 17%), 19% have at most a high school degree or GED (HRS: 53%), 33% reported some college (HRS: 6%), and 33% obtained a four-year college degree or advanced degree (HRS: 24%). Income categories were divided into approximate quintiles in MESA with the highest quintile making $75,000+ and the lowest quintile making $20,000 or less per year. In HRS, the income distribution is skewed slightly more to the right, with 36% of respondents reporting income greater than or equal to $75,000 per year at the time of blood collection. At the time of Exam 3 in 2004-2005, prior to the 2008 housing crisis, the majority of MESA participants (57%) reported having multiple assets that included a home or land, and only 10% reported having zero assets in a home, land, car, or stocks/bonds. In 2016, after the 2008 housing crisis, only 26% of HRS respondents reported multiple assets (including a home), and 15% reported no assets. In terms of lifestyle behaviors, 48% of MESA and 44% of HRS respondents reported never smoking and approximately 10% reported that they were current smokers in both samples. Most participants reported being non-drinkers or drinking fewer than two drinks per day (MESA: 94%; HRS: 90%). In both samples, the majority were overweight or obese, or reported a BMI>25 (MESA: 80%; HRS: 72%).

### Adult socioeconomic disadvantage and epigenetic aging

To capture the multi-dimensional nature of SES and reduce the burden of multiple hypothesis testing, our primary analyses utilized a single SES index that was constructed by taking the unweighted average across all six SES indicators (**Methods**). Higher values of the SES index indicate worse socioeconomic disadvantage, or lower educational attainment and household income, fewer assets in wealth, and greater occupational and neighborhood disadvantage. The SES index is correlated 0.98 in MESA and 0.95 in HRS (all *p*<0.0001) with the first principal component of these SES attributes.

**Table 3** displays the SES index association results for all eight epigenetic clocks in MESA and the HRS (see **Table S5** and **S6** for full results). Model 1 includes controls for age, female sex, race, whether or not the respondent is married, and cohort-specific controls (**Methods**). Model 2 adjusts for lifestyle behaviors that are highly correlated with socioeconomic status and DNAm age (smoking behavior, BMI, and alcohol consumption). For the Horvath 1, Horvath 2, Hannum, Levine, Lin, and GrimAge clocks, a positive coefficient on the SES index variable indicates that an increase in adult socioeconomic disadvantage is associated with an increase in epigenetic aging in years (adjusting for chronological age). For the epiTOC regressions, the coefficient of interest can be interpreted as the percent increase in DNAm at sites with age-related cell-replication errors for every one-point increase in the SES index. DunedinPoAm coefficients represent the rate of aging in years relative to a one-point increase in the SES index. To facilitate comparison across clocks, **Figure 1** also provides coefficient estimates for Model 1 and Model 2 that are standardized and interpretable as Pearson r effect sizes.

**Figure 1.**
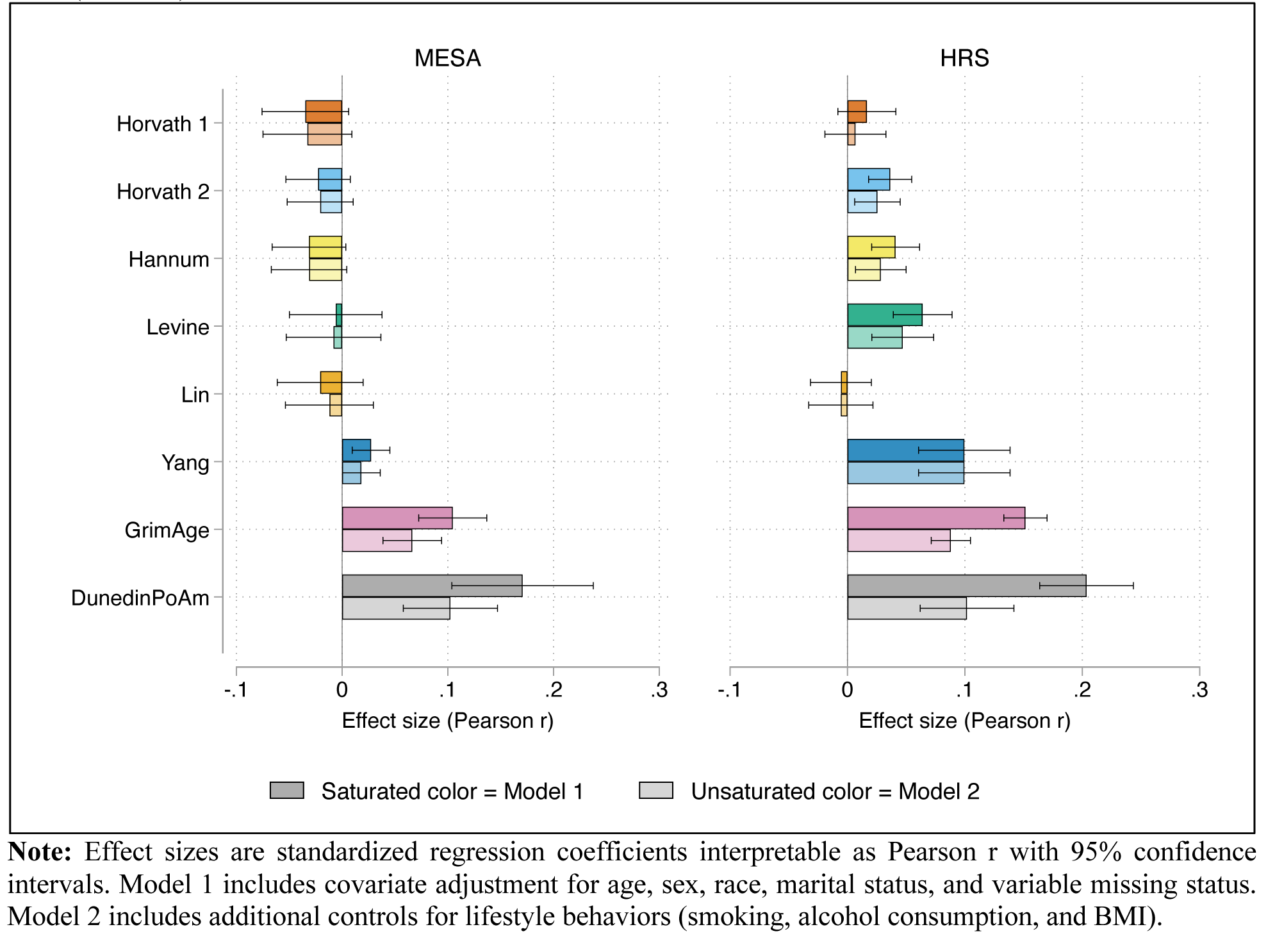
Effect size (Pearson r) estimates for SES Index-Clock associations in MESA (n=1,211) and HRS (n=4,018)

**Table 3.**
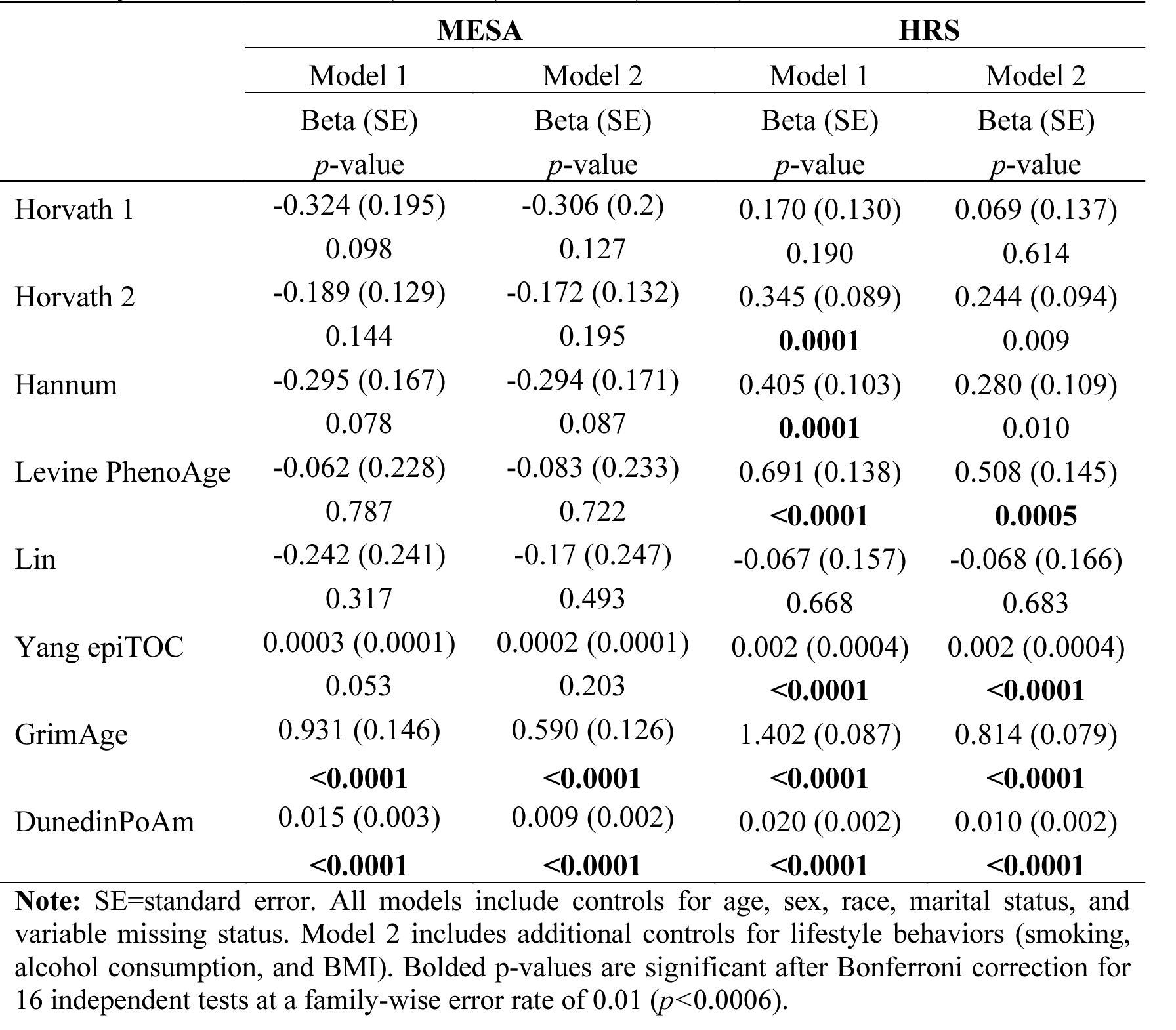
Association between epigenetic clocks and SES index with and without adjustment for lifestyle behaviors, MESA (n=1,211) and HRS (n=4,018)

In both the HRS and MESA, the GrimAge and Dunedin PoAm clocks are positively and significantly associated with the SES index (Bonferroni corrected *p*-value<0.01). For the GrimAge clock, a one-point increase in the SES index is associated with a 0.93-year increase in epigenetic aging in MESA and a 1.4-year increase in HRS. For the PoAm clock, a one-point increase in the SES disadvantage index is associated with accelerated aging at a rate of approximately 0.02 years, or 0.24 months per calendar year for both the MESA and HRS samples. To put this into perspective, the GrimAge estimate in the HRS indicates that an individual with an SES index of five who has the worst socioeconomic standing is, on average, 5.6 years older biologically than an individual with an SES index of one. SES–GrimAge and SES–DunedinPoAm effect size estimates are also higher in magnitude compared to other clock estimates (prior to adjusting for lifestyle behaviors) (**Figure 1**).

Adding lifestyle factors to the analyses does not change the overall significance of these associations, but we do see partial mediation of the SES index–clock relationship: in MESA and the HRS, the SES index coefficient drops by approximately 37%-42% for GrimAge and by 40%- 50% for DunedinPoAm. In MESA, current or former smoking status is associated with higher GrimAge and DunedinPoAm epigenetic aging (**Table S5**). In HRS, smoking status, consuming more than two alcohol beverages per day, and obesity (BMI≥30) are all associated with higher GrimAge and DunedinPoAm measures relative to the reference groups (never smokers, non-drinkers, and BMI≤25) (**Table S6**).

In the HRS, we also see significant associations between the SES index and the Horvath 2, Hannum, Levine, and Yang clocks. However, after adjusting for lifestyle behaviors, the Horvath 2 and Hannum associations are not significant after Bonferroni correction. In MESA, the remaining six clocks display insignificant associations, and with the exception of the Yang clock, these associations are negative. We hypothesize that certain clock associations may not replicate in MESA due to the smaller sample size, larger proportion of individuals from diverse ancestral groups, and/or the fact that DNAm was profiled in one cell type (monocytes). Because MESA participants were required to be without clinically apparent cardiovascular disease (CVD) when they were recruited, it is also possible that MESA participants were healthier at baseline than HRS participants. In supplemental analyses, we show that the estimates in **Table 3** are not driven by MESA exam site (**Table S7**), longitudinal timing of when SES questions were asked relative to blood collection (**Table S8**), or cell proportion adjustment (in HRS) (**Table S9**).

Overall, the SES index explains at most ∼3% of the variance in epigenetic clock outcomes (for DunedinPoAm in the HRS sample), and after adjusting for lifestyle behaviors, the proportion of clock variance explained by the SES index falls to less than 1% for the Levine, Yang, GrimAge, and DunedinPoAm clocks (**Figure S1**). This suggests that although several clocks display fairly robust associations with socioeconomic disadvantage at older ages, social forces appear to explain a relatively small proportion of the epigenetic gradient in aging across these clocks.

### Associations between SES dimensions and epigenetic clocks

To assess whether specific dimensions of socioeconomic disadvantage are driving the relationships reported in **Table 3 and Figure 1**, we examined associations between the clocks and each component of the SES index if a clock displayed significant associations with the full SES index. We regressed the epigenetic clocks on each SES dimension separately with and without controls for lifestyle behaviors (i.e., adjusting for either Model 1 or Model 2 covariates). To facilitate comparison across SES dimensions, we standardized regression coefficients so that they are interpretable as Pearson r effect sizes. Results from Model 2 for the Levine, Yang, GrimAge, and DunedinPoAm clocks are presented in **Figure 2** for the HRS sample. In MESA, results for DunedinPoAm and GrimAge closely parallel those in the HRS for education, income, occupation, and neighborhood environment, both in magnitude and significance (unstandardized results for both samples are presented in **Tables S10-S11**).

**Figure 2.**
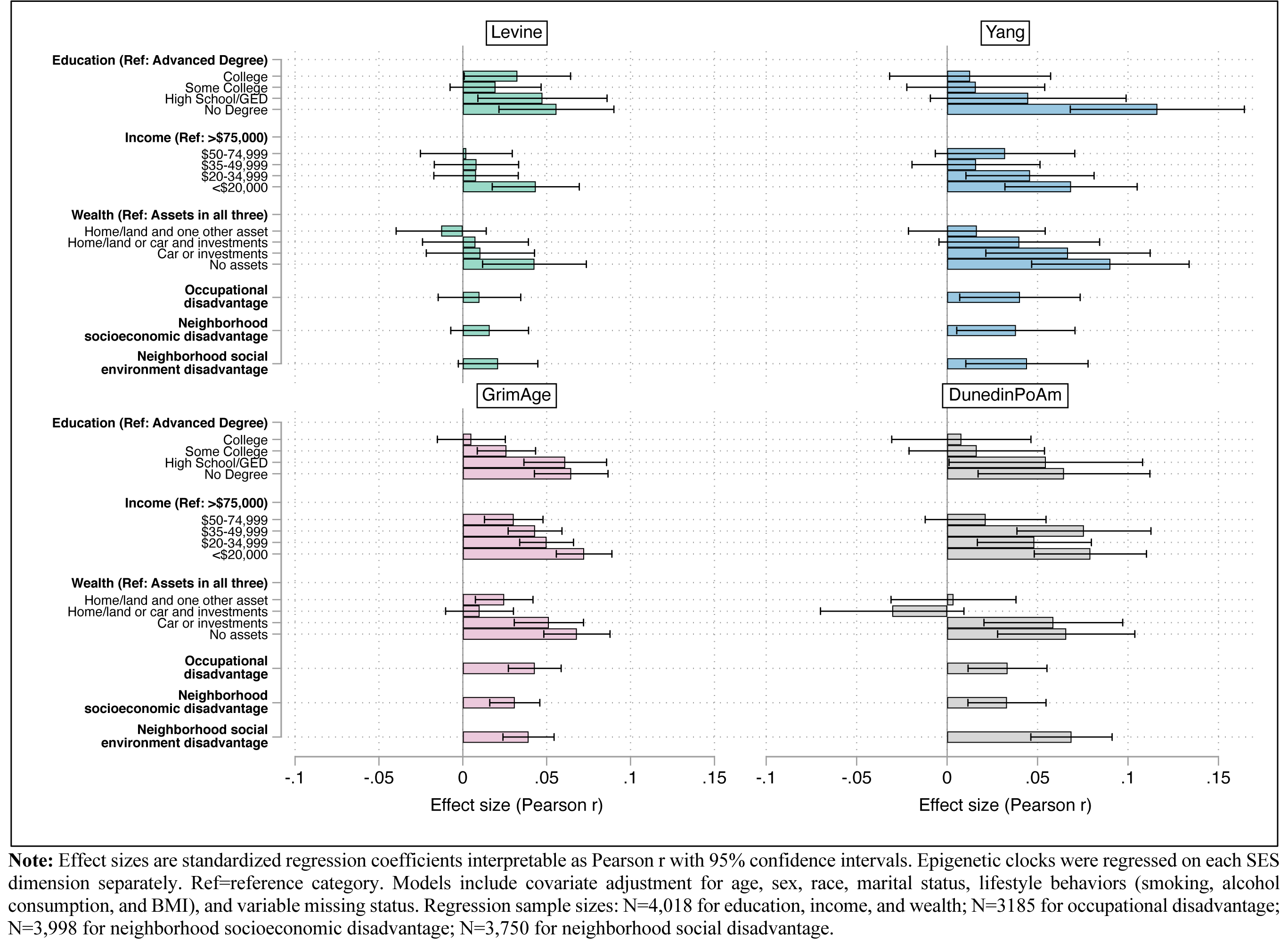
Effect size (Pearson r) estimates for associations between Levine, Yang, GrimAge, and Dunedin PoAm clocks and components of the SES index, HRS

Several patterns emerge. First, across all clocks, we see non-zero coefficient estimates for educational attainment, household income, and household wealth, particularly for the most disadvantaged groups. Individuals with no degree are epigenetically older than individuals with an advanced degree; individuals making <$20,000 per year are aging epigenetically faster than individuals making $75,000 per year; and individuals with no assets in wealth are aging faster than individuals who own land, a home, a car, and/or have financial investments. For the GrimAge and DunedinPoAm clocks, we also see significantly higher epigenetic age for individuals with no college degree, less than $50,000 in household income, and fewer assets in wealth.

Secondly, for the GrimAge, DunedinPoAm, and Yang epiTOC clocks, additional positive and statistically significant relationships emerged for occupational disadvantage and neighborhood-level socioeconomic and social disadvantage. Higher values of the occupational disadvantage index indicate that an individual worked in an occupation that was ranked as being less prestigious in a representative sample of U.S. workers (**Methods**). Higher neighborhood socioeconomic and social disadvantage scores are Census tract-based metrics of socioeconomic status and aesthetic quality, safety, and social cohesion, respectively, that have been previously linked with the DNAm of stress- and inflammation-related genes in MESA (**Methods**).^69^

Third, if we compare coefficient estimates across Models 1 and 2, with the exception of GrimAge, SES coefficients in models adjusted for smoking, BMI, and alcohol use are not significantly different from unadjusted estimates (**Tables S10-S11)**. Conversely, GrimAge estimates that adjust for lifestyle behaviors are significantly lower in magnitude for education (no degree), income (<$35,000), occupation, and neighborhood socioeconomic disadvantage. Since GrimAge is in part trained on smoking-pack-years, it is perhaps unsurprising that a significant proportion of the SES-epigenetic age relationship is explained by smoking and related health behaviors. However, even in the case of GrimAge, lifestyle does not fully mediate the SES gradient, which suggests that differences in health behaviors alone do not drive differences in epigenetic aging across social classes.

Overall, whether or not we adjusted for lifestyle behaviors, no one dimension of SES stands out—i.e., effect size estimates are comparable across models. To examine this further, in supplementary analyses, we tested additional models in the HRS that adjusted simultaneously for all six SES components (**Table S12**). Across all four clocks, significant relationships for educational attainment and income continue to hold, and for GrimAge and DunedinPoAm clocks, significant relationships for wealth also persist.

### Childhood socioeconomic disadvantage and epigenetic aging

Several studies have found that exposure to trauma or socioeconomic disadvantage in early life is associated with accelerated epigenetic aging,^10, 41, 47, 49, 50, 55, 57, 59, 70^ particularly during critical periods of development.^55, 70^ The vast majority of these studies have assessed associations with first generation clocks. In some cases, childhood SES was found to explain or fully mediate relationships between adult SES and the Horvath and Hannum clocks, which suggests that adult SES may be acting as a proxy for unmeasured conditions earlier in life.^41^

To assess whether childhood socioeconomic position is driving the findings we report for adult SES, we added mother’s education to the fully adjusted SES index model in the HRS sample (higher values equal more years of education). Parental education is seen as an important determinant of social mobility, and past studies have shown that compared to other social background characteristics, parental education exerts the strongest influence on a child’s own educational attainment.^71–73^ Including father’s education in addition to mother’s education did not appear to add any additional explanatory power and the coefficients for father’s education were null or marginally significant in both samples (**Table S13**). **Figure 3** reports standardized coefficients with and without adjustments for mother’s education (full results are available in **Table S13**). Similar to past research, we find a significant relationship between childhood SES and the Hannum clock, but not the Horvath clock, that fully mediates the adult SES relationship (*p*<0.0001). Mother’s education is also significantly associated with the Yang clock (*p*<0.0001), but its inclusion in the model does not significantly alter the magnitude of the adult SES coefficient. Interestingly, we do not find any relationship between mother’s education and the GrimAge or DunedinPoAm clocks, and its addition to the model does not affect the SES coefficient. Thus, particularly for second generation clocks, childhood SES as measured by parental education, does not appear to be acting as a proxy for adult socioeconomic disadvantage.

**Figure 3.**
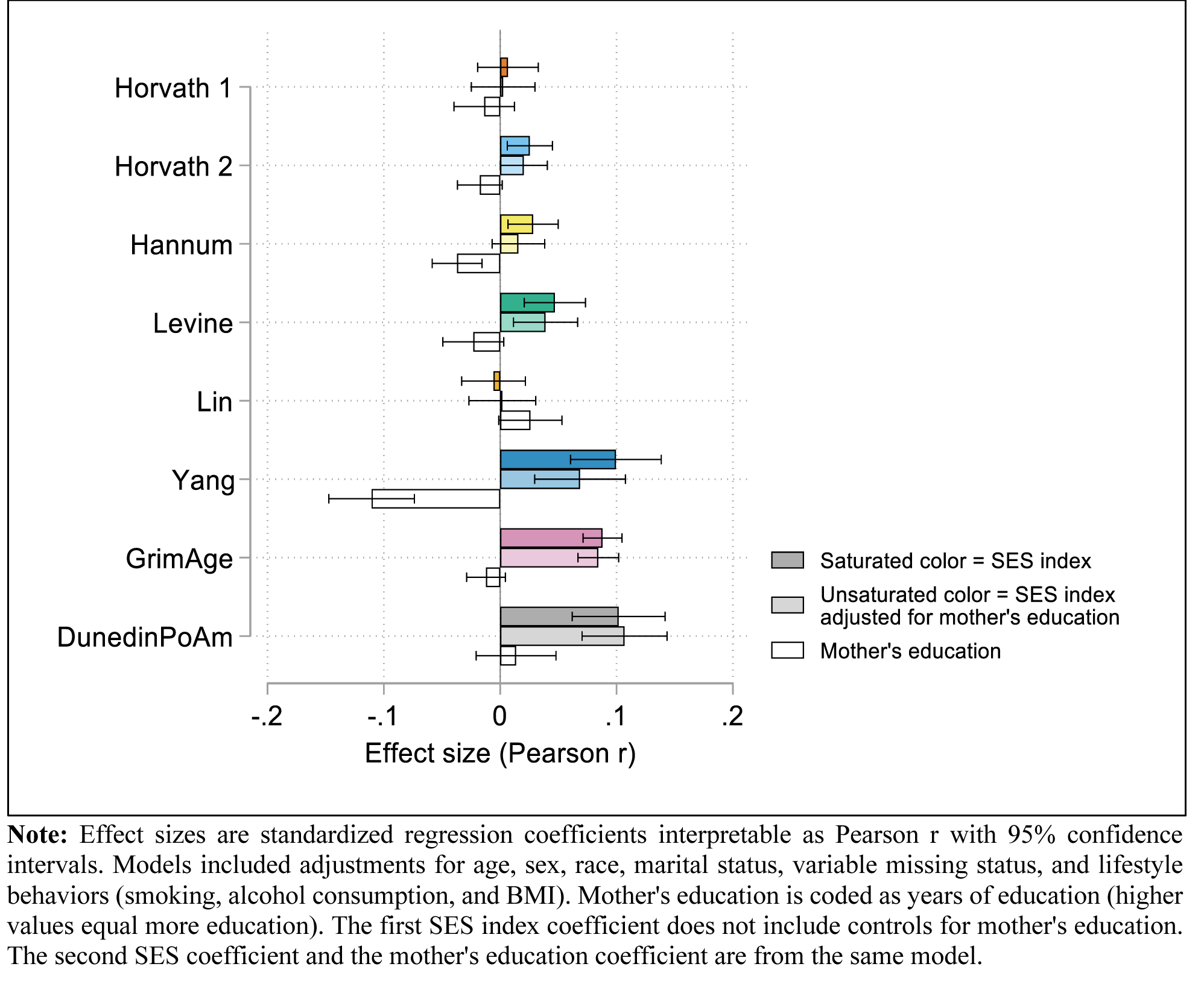
Effect size (Pearson r) estimates for associations between epigenetic clocks and the SES index with and without adjustments for mother’s education, HRS (n=4,018)

### Sensitivity analysis by race, sex, and age

SES-epigenetic clock associations have been shown to differ by sex, age and race.^48, 61, 70^ **Figure 4** displays SES index-clock associations stratified by ancestry, sex, and age, and adjusted for lifestyle behaviors (full results with and without adjustments for lifestyle behaviors are provided in **Tables S14** and **S15**). Clock associations appear to be most significant in European ancestry individuals. However, the GrimAge, DunedinPoAm, and Yang clocks also display significant associations in individuals of Hispanic ethnicity and African ancestry, and overall estimates across ancestral groups are not significantly different from one another. With the exception of stronger GrimAge and DunedinPoAm associations for males in MESA, associations are either slightly stronger for females or do not differ significantly between males and females. Estimates do not appear to differ dramatically by age group; however, in HRS the GrimAge and DunedinPoAm estimates are higher for individuals <65 years old and differ significantly from individuals aged 65+. In the case of DunedinPoAm, this could in part reflect the fact that the clock was trained on biomarkers that were assessed in the Dunedin Study when respondents were between the ages of 26 and 38. Because the clock was trained to capture rate of aging in midlife, it may be picking up on DNAm changes related to extreme aging differences in accelerated outliers who are less likely to survive into older ages.

**Figure 4.**
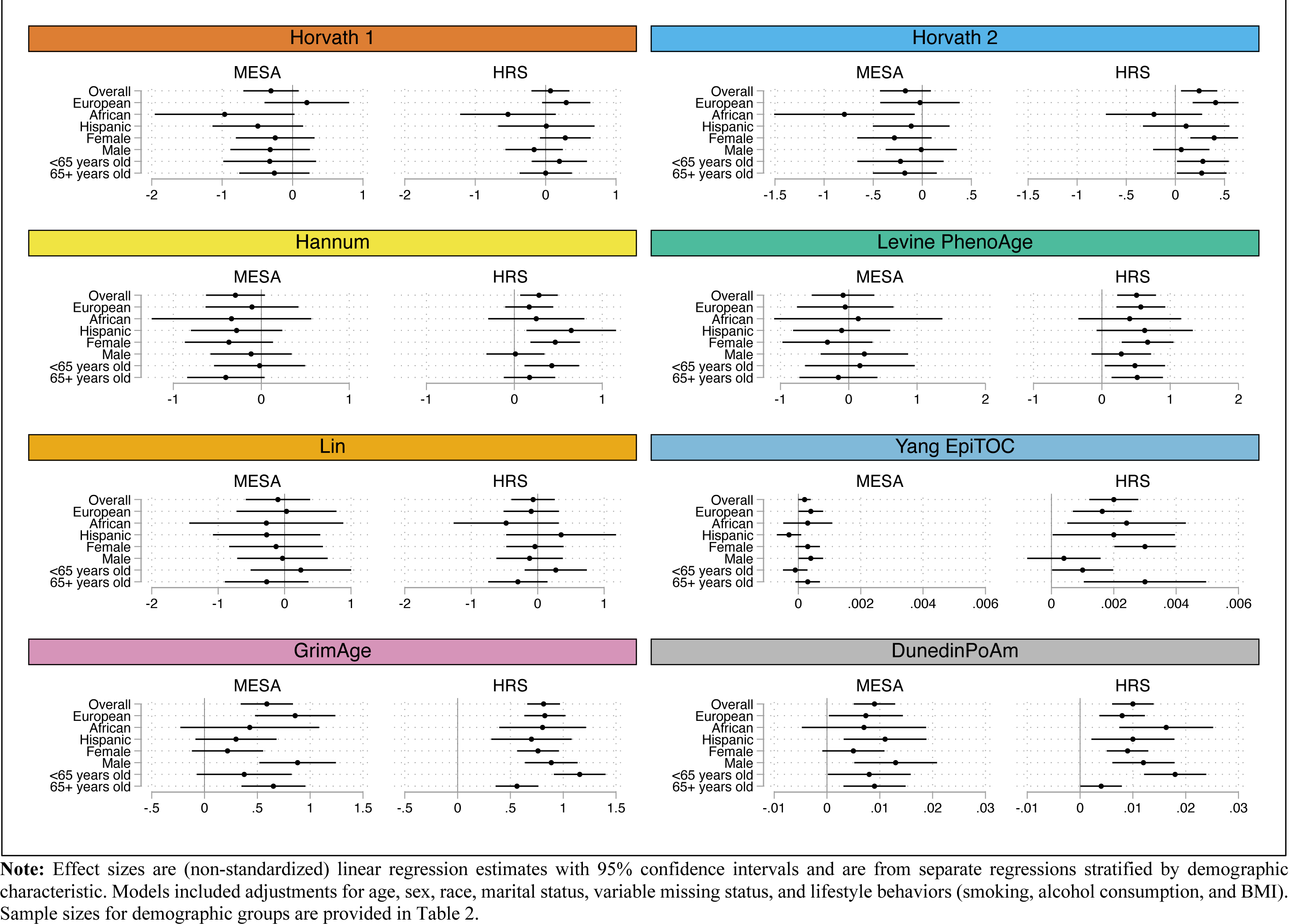
Forest plots of the association between epigenetic clocks and the SES index by demographic characteristic, MESA and HRS

### Genetic moderation of SES-clock associations

Published GWAS of DNAm age have examined single nucleotide polymorphism (SNP)-level associations with IEAA and EEAA measures (i.e., the Horvath 1 and Hannum clocks) in individuals of European ancestry.^66, 74^ The IEAA and EEAA terminology has been used to refer to the observation that clocks trained in whole blood may be more reflective of age-related changes in leukocyte composition, whereas multi-tissue clocks are independent of these changes. For example, multi-tissue epigenetic clocks like the Horvath 1 clock are said to measure (cell-)intrinsic aging because they are largely independent of cell composition changes, whereas the Hannum and Levine clocks, for example, also measure extrinsic aging.^39^ IEAA measures adjust Horvath 1 for cell composition, while EEAA measures incorporate cell composition with the Hannum clock to further capture changes related to aging in white blood cells. Heritability estimates of IEAA and EEAA range from approximately 10%-19% for SNP-based heritability estimates,^66, 68^ to approximately 40% for pedigree-based estimates .^39, 74^

Given that epigenetic aging is to an extent regulated by genetics, we examined whether SES-clock relationships are moderated by polygenic scores (PGSs) for IEAA and EEAA. PGSs were constructed in the MESA and HRS European ancestry samples using weights from the most recent GWAS of DNAm age (**Methods**).^66^ To optimize the predictive performance of the PGS, we utilized a novel PGS-tuning method that selects the optimal p-value threshold using GWAS summary statistics as opposed to within sample validation, which may result in overfitting.^61^ Since GWAS estimates are currently only available for the Horvath 1 and Hannum clocks, we were not able to construct PGSs that capture the unique genetic underpinnings of all eight clocks. However, to assess any potential genetic overlap between the clocks, we assessed the predictive performance of the IEAA and EEAA PGSs across all eight clocks (**Tables S16** and **S17**). As expected, the IEAA PGS is strongly associated with the Horvath 1 clock in MESA (*p*=1.84E-09) and HRS (*p*=5.93E-08). We also find significant associations between the IEAA PGS and the Levine clock in MESA (*p*=0.0001) and HRS (*p*=0.001). We do not see a significant association between the EEAA PGS and any of the clocks in MESA, most likely because changes in cell proportions will not be captured when DNAm is profiled in one cell type. Conversely, in the HRS, we find EEAA PGS associations with the Hannum (*p*=1.46E-08), Horvath 2 (*p*=7.63E-05), and Horvath 1 (*p*=0.001) clocks.

**Table 4** reports results from regressions that add the PGS and its interaction with the SES index to the SES index-clock analysis for clocks that were significantly correlated with the IEAA PGS in one or both samples (**Table S18** reports EEAA PGS results in HRS). Apart from the Hannum clock in HRS, the IEAA PGS is significantly associated with the Horvath 1 and Levine clocks in the SES model. The magnitude of the PGS associations are also significantly higher than the SES index associations, indicating a strong independent effect of genetic predisposition. Because higher values of the PGS are associated with accelerated epigenetic aging, this suggests that genetically-at-risk individuals from disadvantaged social backgrounds may experience more rapid declines in health than low-risk individuals from comparable backgrounds. Genetic predisposition does not appear to moderate the relationship between the SES index and the clocks; however, this may in part be due to the relatively weak SES correlations with the Horvath and Hannum clocks and/or lack of statistical power to detect moderation effects. Thus, as more epigenetic data and larger GWAS become available, additional research is needed to assess the degree to which genetic predisposition moderates the relationship between socioeconomic disadvantage and accelerated epigenetic aging (particularly in second-generation clocks).

**Table 4.** Genetic moderation of the SES-epigenetic clock relationship by intrinsic epigenetic age acceleration (IEAA) polygenic score (PGS), European ancestry subsample, MESA (n=582) and HRS (n=2,265)

|  | Horvath 1 |  | Hannum |  | Levine |  |
| --- | --- | --- | --- | --- | --- | --- |
|  | MESA | HRS | MESA | HRS | MESA | HRS |
|  | Beta (SE) | Beta (SE) | Beta (SE) | Beta (SE) | Beta (SE) | Beta (SE) |
|  | *p*-value | *p*-value | *p*-value | *p*-value | *p*-value | *p*-value |
| SES Index (std) | 0.128 (0.233) | 0.195 (0.183) | -0.193 (0.209) | 0.165 (0.145) | -0.245 (0.278) | 0.428 (0.188) |
|  | 0.583 | 0.288 | 0.358 | 0.256 | 0.380 | **0.023** |
| PGS (std) | 1.231 (0.203) | 0.759 (0.144) | 0.686 (0.183) | 0.127 (0.114) | 0.976 (0.242) | 0.504 (0.148) |
|  | **<0.0001** | **<0.0001** | **0.0002** | 0.265 | **<0.0001** | **0.0007** |
| SES Index*PGS | -0.201(0.213) | -0.016 (0.150) | 0.15 (0.192) | -0.219 (0.119) | -0.053 (0.255) | 0.159 (0.154) |
|  | 0.345 | 0.915 | 0.434 | 0.065 | 0.835 | 0.303 |

**Note:** SE=standard error; SES=socioeconomic status; std=standardized. All models were estimated using linear regression and include controls for age, sex, race, marital status, variable missing status, lifestyle behaviors (smoking, alcohol consumption, and BMI), and the first 10 principal components of the genetic data matrix. SES index and polygenic scores were standardized to have a mean of 0 and a standard deviation of 1.

## Discussion

A better understanding of the relationship between SES and epigenetic aging may lend insight into the specific social and behavioral mechanisms that underpin the SES-mortality gradient. In this study, we found robust associations between a multi-dimensional SES index and the GrimAge and DunedinPoAm epigenetic clock algorithms in two U.S. aging studies—MESA and the HRS. In the HRS, the larger of the two samples, associations between SES and the Levine PhenoAge and Yang epiTOC clocks were also evident. Across the separate dimensions of the SES index, educational attainment and income displayed the most robust associations with the Levine, Yang, GrimAge, and DunedinPoAm clocks, particularly for individuals in the most disadvantaged categories (i.e., individuals with no degree or at most a GED/HS degree and individuals with <$35,000 per year in household income). The GrimAge and DunedinPoAm clock associations were partially (but not fully) mediated by the inclusion of lifestyle behaviors, which suggests that differences in health behaviors alone cannot explain the SES gradient in epigenetic aging. At most, the SES index explains ∼3% of the variation in epigenetic aging, and ∼1% after accounting for lifestyle factors. In a smaller subsample of European ancestry individuals, genetic risk for accelerated intrinsic epigenetic aging (as measured by a PGS) is independently associated with higher Horvath, Levine, and Hannum EAA, signifying that genetic risk and social disadvantage may contribute additively to faster biological aging. Overall, our results suggest that certain clocks may be more useful for informing research on the social determinants of health and aging. In addition, as the availability of epigenetic and genetic profiles in population studies increases, further exploration of the feedback between the social environment and genetic mechanisms that impact epigenetic regulation may be informative for health disparities research.

Several open questions remain. In particular, in this study we cannot tease apart why some clocks are more correlated with SES than others. One hypothesis supported by previous research suggests that there are distinct pathways of epigenetic aging across aging tissues and cell states, and that pathways of epigenetic aging captured in each of the clocks may vary according to the social and environmental exposures of the particular sample(s) and the outcome measures that were used to train the clocks.^68^ For example, the Levine and DunedinPoAm clocks were trained in study populations in Italy and New Zealand. These cohorts were exposed to different cultural norms, social policies, and historical conditions that may have differentially affected social pathways of epigenetic aging.^10^ Accordingly, to capture distinct social pathways of epigenetic aging, it may be useful to train epigenetic predictors that can more directly capture social gradients in health and mortality, either by using larger, more socioeconomically diverse populations to train clocks in distinct samples or by training clocks directly on socioeconomic outcomes. In line with a recent approach carried out by Liu et al. (2020), it may also be useful to deconstruct existing clocks into distinct, nonoverlapping submodules that are highly associated with SES at the CpG level and then recombine the most robust signals into a single, novel clock.

This study has several strengths. We replicated our findings in two large aging studies, used multiple measures of SES, examined relationships with first- and second-generation epigenetic clocks, and explored the extent to which current measures of genetic risk affect our findings. Several limitations should also be mentioned. Most importantly, we were not able to draw conclusions regarding the causal relationship between epigenetic clock measures and socioeconomic position in childhood or adulthood. Minimal research exists on causal relationships and should be further investigated in future studies. Additionally, although our results display a high degree of internal validity, they may not replicate in other cohorts with divergent socioeconomic exposures or demographic characteristics. We also acknowledge that the epigenome is highly sensitive to disease and various environmental exposures that may have affected DNAm at the time of blood collection and that may be confounding the associations we report. Thus, future research that incorporates longitudinal measures of biological, behavioral, and social factors into the same framework may help solidify linkages between these factors while also assessing their relative contribution to individual differences in aging.

## Methods

### Multi-Ethnic Study of Atherosclerosis (MESA)

MESA is a population-based longitudinal study designed to identify risk factors for the progression of subclinical cardiovascular disease (CVD).^75^ A total of 6,814 non-Hispanic white, African-American, Hispanic, and Chinese-American women and men aged 45–84 without clinically apparent CVD were recruited between July 2000 and August 2002 from the following 6 regions in the US: Forsyth County, NC; Northern Manhattan and the Bronx, NY; Baltimore City and Baltimore County, MD; St. Paul, MN; Chicago, IL; and Los Angeles County, CA. Each field center used locally available sources to recruit participants, including lists of residents, lists of dwellings, and telephone exchanges. During MESA Exam 5 (between April 2010 and February 2012), DNA methylation was assessed on a random subsample of 1,264 non-Hispanic white, African American, and Hispanic MESA participants aged 55–94 years old from the Baltimore, Forsyth County, New York, and St. Paul field centers who agreed to participate in an ancillary study examining the effects of methylation on CVD. We excluded 53 respondents with exposure data. Our final sample consisted of 1,211 MESA participants with genome-wide genetic data and DNAm data from monocytes at Exam 5. Genotype and phenotype information for MESA were obtained from the National Center for Biotechnology Information’s database of Genotypes and Phenotypes (NCBI dbGaP study accession: phs000209.v11.p3 - MESA SNP Health Association Resource (SHARe)). This study was approved by the Institutional Review Boards of all MESA field centers, the MESA Coordinating Center, the University of Wisconsin-Madison, and the University of Michigan.

### Health and Retirement Study (HRS)

The HRS is a nationally representative, longitudinal panel study of individuals over the age of 50 and their spouses. The study is sponsored by the National Institute on Aging (NIA U01AG009740) and is conducted by the University of Michigan.^76, 77^ Launched in 1992, the HRS introduces a new cohort of participants every six years and interviews around 20,000 participants every two years. To maximize sample size, we compiled data from 13 waves (1992-2016). We used data from the RAND HRS Longitudinal File 2016 (V2) for all longitudinal demographic and socioeconomic data as well as data on health behaviors.^78^ Neighborhood social disadvantage data on neighborhood disorder and neighborhood social cohesion were taken from the 2006-2016 Psychosocial and Lifestyle Questionnaire (PLQ)^79^ and merged to respondents at the Census tract level using restricted data on location of primary residence from the 2006-2016 waves. Neighborhood socioeconomic disadvantage measures were constructed using data from the restricted HRS Contextual Data Resource (HRS-CDR), which contains data from the 2005-2018 American Community Survey (ACS) linked to respondents at the Census tract level.^80^ Restricted three-digit Census occupation codes from the 1992-2016 HRS waves were used to construct occupational prestige measures. Access to HRS restricted data was approved by HRS and the University of Wisconsin-Madison Institutional Review Board.

DNA methylation data was collected as part of the 2016 HRS Venous Blood Study. A total of 4,018 samples passed QC. The DNAm sample is racially and socioeconomically diverse and representative of the full HRS sample.^81^ We utilized all 4,018 respondents in the DNAm sample in our SES-clock analysis. In sensitivity analyses, we adjusted for cell-type proportions using results from a white blood cell (WBC) differential assay.^82^ Genotype data on ∼15,000 HRS participants was collected from a random subset of the ∼26,000 total participants that were selected to participate in enhanced face-to-face interviews and saliva specimen collection for DNA in 2006, 2008, and 2010. Imputed HRS genotype data were accessed through dbGap (phs000428). For the PGS analysis, we restricted our sample to 2,265 European ancestry individuals with DNAm and genotype data.

### MESA DNA methylation data

A detailed description of the data extraction and processing procedures used for DNA methylation can be found in Liu et al. (2013). Briefly, blood was drawn in the morning after a 12 hour fast. Monocytes were isolated using AutoMACs automated magnetic separation units (Miltenyi Biotec, Bergisch Gladbach, Germany) and were consistently >90% pure. Samples were plated using a stratified random sampling technique to reduce bias due to batch, chip, and position effects. Methylation was measured using the Illumina HumanMethylation450 BeadChip, and bead-level data were summarized in GenomeStudio. Quantile normalization was performed using the lumi package with default settings.^84^ Quality control measures included checks for sex and race/ethnicity mismatches and outlier identification by multidimensional scaling plots. Criteria for elimination included: ‘detected’ methylation levels in <90% of MESA samples (using a detection p-value threshold of 0.05) or overlap with a non-unique region. 65 probes that assay highly polymorphic single nucleotide polymorphisms (SNPs) rather than methylation were also excluded.^85^ The methylation level for each site was computed as the M-value, the log ratio of the methylated to the unmethylated signal intensity.^86^ We adjusted for chip and position effects in all analysis.

### HRS DNA methylation data

Detailed information on the 2016 Venous Blood Study is provided in the VBS 2016 Data Description.^82^ Blood was collected from consenting respondents during in home phlebotomy visits. Every attempt was made to schedule the blood draw within four weeks of the 2016 HRS core interview. Fasting was recommended but not required. Methylation was measured using the Infinium Methylation EPIC BeadChip. Samples were randomized across plates by key demographic variables (i.e., age, cohort, sex, education, race/ethnicity) with 40 pairs of blinded duplicates.^81^ Analysis of duplicate samples showed a correlation >0.97 for all CpG sites. The *minfi* package in R software was used for data preprocessing, and quality control. 3.4% of the methylation probes (n=29,431 out of 866,091) were removed from the final dataset due to suboptimal performance (using a detection p-value threshold of 0.01). Analysis for detection p-value failed samples was done after removal of detection p-value failed probes. Using a 5% cut- off (*minfi*) 58 samples were removed. Sex mismatched samples and any controls (cell lines, blinded duplicates) were also removed. High quality methylation data is available for 97.9% samples (n=4,018). Missing beta methylation values were imputed with the mean beta methylation value of the given probe across all samples prior to constructing DNAm age measures.

### MESA genotype data

DNA samples were genotyped in blood samples using the Affymetrix 6.0 SNP array. Samples with call rate <95%, duplicated samples, and those with gender mismatch were excluded. Further details are described elsewhere.^75^ Genotyped data were imputed to the 1000 Genomes Project cosmopolitan reference panel phase 1 (version 3) using IMPUTE2.^87^ SNPs with low imputation quality (info<0.8) were excluded. MESA was stratified into four ethnic-specific samples, which were verified via clustering in EIGENSTRAT.^88^

### HRS genotype data

Genotyping was conducted by the Center for Inherited Disease Research (CIDR) in 2011, 2012, and 2015 (RC2 AG0336495, RC4 AG039029). Full quality control details can be found in the Quality Control Report. Genotype data on over 15,000 HRS participants was obtained using the llumina HumanOmni2.5 BeadChips (HumanOmni2.5-4v1, HumanOmni2.5- 8v1), which measures ∼2.4 million SNPs. Genotyping quality control was performed by the Genetics Coordinating Center at the University of Washington, Seattle, WA. Individuals with missing call rates >2%, SNPs with call rates <98%, HWE p-value < 0.0001, chromosomal anomalies, and first-degree relatives in the HRS were removed. Imputation to 1000G Phase I v3 (released March 2012) was performed using SHAPEIT2 followed by IMPUTE2. The worldwide reference panel of all 1,092 samples from the Phase I integrated variant set was used. These imputation analyses were performed and documented by the Genetics Coordinating Center at the University of Washington, Seattle, WA. All positions and names are aligned to build GRCh37/hg19.

### Epigenetic clock outcomes

We used measures of DNAm age from eight existing epigenetic clocks listed in Table 2. In MESA, we constructed these clocks from CpG-level data. In HRS, we used publicly available clocks constructed by the HRS.^81^ In both studies, DNAm age was computed as the weighted sum of age-related CpGs, where weights were defined using a penalized regression model. **Table 1** details the number of CpG sites used to compute each epigenetic clock according to author-specific algorithms. In regression analyses, we adjusted for age so that the dependent variable represents epigenetic age net of chronological age.

### Education

In both studies, education was measured using dichotomous variables for highest degree obtained. Responses were categorized as “no degree”, “HS degree or GED”, “some college” (i.e., less than a bachelor’s degree or an associate degree), “college” (bachelor’s degree), or “advanced degree” (i.e., masters/MBA, JD, MD or PhD degrees).

### Household income

In MESA, respondents selected their household’s income bracket from a series of unfolding brackets rather than reporting a continuous value. We collapsed all 15 income brackets into approximate household income quintiles for the MESA sample as follows: <$5,000-$19,999, $20-34,999, $35,000-$49,000, $50,000-74,999, and ≥$75,000. We used the same income quintile cut points to construct comparable household income categories for HRS participants from continuous, self-reported household income data. In both studies, gross household income includes income from respondent and spouse earnings, pensions and annuities, Social Security retirement, unemployment and workers compensation, other government transfers, and household capital income. Income is reported in 2010 dollars. For MESA, we used income data from Wave 5; for HRS, income data was taken from the 2016 wave.

### Household wealth

MESA data on household wealth is available in Exam 3. Respondents were asked whether they have any assets in a home, in land, in a car, or in financial investments. Financial investments included assets in stocks, bonds, mutual funds, or retirement investments. We used this information to create five mutually exclusive, dichotomous wealth categories: 1) respondent has assets in a home/land, in a car, and in financial investments; 2) respondent has assets in home/land and one other asset (cars or financial investments); 3) respondent has assets in home/land only or has two other assets (car and financial investments); 4) respondent has assets in a car or in financial investments but does not have assets in home/land; and 5) respondent has no assets. For comparability with MESA, we used self-reported data on household assets to construct the same categories for HRS respondents using data from the 2016 wave.

### Occupational disadvantage

We merged occupational prestige scores from the General Social Survey (GSS) to three-digit 1980 or 2000 Census occupation codes in both MESA and the HRS to capture the degree of status associated with a given occupation. To construct a measure of occupational disadvantage, scores were standardized to have a mean of 0 and a standard deviation of 1 and then reverse coded so that higher values reflect less prestige. We used the average occupational disadvantage score across all available exams/waves that MESA/HRS respondents reported working. Occupation data were available for MESA exams 1-3 and across all waves in the HRS.

Occupational prestige scores are weighted average ranks of occupations by samples of workers that have been found to be consistent across raters from different social positions and contexts over time,^89–92^ and have been found to capture aspects of occupational status related to health and health behaviors that are independent of education and income.^93, 94^ We used data on occupational prestige from the 2012 GSS.^89^ A sample of ∼1,000 individuals rated 90 occupations each; a rotation of occupation ratings across respondents resulted in ratings for 860 occupational titles. Respondents were asked to rate the social standing of occupations on a hypothetical ladder from top to bottom. Scores adjusted for reviewer-specific effects so that they reflect occupational differences rather than differences between raters using hierarchical linear modeling (HLM).

### Neighborhood characteristics

We follow past work with MESA that deployed neighborhood-based scores for socioeconomic disadvantage and the social environment created at the Census tract level (where the Census tract is used as a proxy for neighborhoods).^95^ In MESA, the **neighborhood socioeconomic disadvantage score** for each neighborhood was created based on a factor analysis of 16 census tract level variables from the 2000 Census that reflect dimensions of education, occupation, income, wealth, poverty, employment, and housing. The neighborhood socioeconomic disadvantage score is the weighted sum of the following six standardized variables, which accounted for 49% of the variance in MESA (52% in the HRS) and loaded onto the first factor: (1) percent in census tract with a bachelor’s degree; (2) percent with a managerial/professional occupation; (3) percent with a high school education; (4) median home value; (5) median household income; and (6) percent with household income greater than $50,000 per year. Variables were reverse coded so that higher values on the scale indicate greater neighborhood socioeconomic disadvantage. We used the same methodology to create the neighborhood socioeconomic disadvantage score in the HRS using HRS-linked Census data from the HRS-CDR database.^80^

The **neighborhood social disadvantage score** is the sum of conditional empirical Bayes estimate (CEB) scales for aesthetic quality, safety, and social cohesion. The CEB estimates are more reliable than the census-tract crude means because they borrow information from other census tracts in cases where the sample size per tract is very small. Detailed information on the development and validation of this score in MESA is provided elsewhere.^96^ Briefly, respondents were asked to report their levels of agreement on a 5-point scale (1=strongly agree to 5=strongly disagree) to statements pertaining to neighborhood aesthetic quality, safety, and social cohesion. The CEB estimate for each dimension of the neighborhood social disadvantage score (aesthetic quality, safety, and social cohesion) was estimated from a HLM to account for the fact that each series of questions used to estimate a particular dimension is nested within individuals who are nested within census tracts. The HLM estimates also adjust for site (MESA) or state (HRS) fixed effects, participant age, and sex. The CEB estimates for each dimension were standardized and then aggregated to create the neighborhood social disadvantage score at each MESA/HRS exam/wave. In MESA, the statements for aesthetic quality were: (1) there is a lot of trash and litter on the street in my neighborhood; (2) there is a lot of noise in my neighborhood; (3) my neighborhood is attractive. The statements for safety were: (1) I feel safe walking in my neighborhood day or night; and (2) violence is a problem in my neighborhood. The statements for social cohesion were: (1) people around here are willing to help their neighbors; (2) people in my neighborhood generally get along with each other; (3) people in my neighborhood can be trusted; and (4) people in my neighborhood share the same values. Where necessary, statements were reverse coded so that higher values indicate worse neighborhood social environment (i.e., lower aesthetic quality, less safety, or lower social cohesion).

In the HRS, we used participant evaluations of their neighborhood environment in the 2006-2016 PLQ that probe how respondents feel about their local area (i.e., everywhere within a 20-minute walk or within a mile of their home).^79^ Participants were asked to mark boxes numbered from one to seven on a line, where seven indicated they agree more strongly with the statement. In the HRS PLQ, statements for aesthetic quality were: (1) vandalism and graffiti are a big problem in this area; (2) this area is always full of rubbish and litter; and (3) there are many vacant or deserted houses or storefronts in this area. The statement for safety was (only one measure was available): (1) people would be afraid to walk alone in the area after dark. The statements for social cohesion were: (1) most people in this area can’t be trusted; (2) most people in this area are unfriendly; (3) if you were in trouble, there is nobody in this area who would help you; and (4) I feel that I don’t belong to this area.

We used responses from the entire HRS sample to construct census-tract level estimates of neighborhood socioeconomic and social disadvantage for participants with DNAm data. For both neighborhood scores, we used the cumulative average across all exams/waves to capture average exposure. Scores were standardized to have a mean of 0 and standard deviation of 1 for all analysis.

### Socioeconomic status (SES) index

To capture the multi-dimensional nature of SES and reduce the burden of multiple hypothesis testing, we constructed an SES index by taking the (unweighted) average across all six SES indicators. For each indicator, we ranked outcomes on a scale from 1 to 5 so that higher values equal more disadvantage as follows: (a) <u>Education</u>: 1=advanced degree, 2=college, 3=some college, 4=HS degree/GED, 5=no degree; (b) <u>Household income</u>: 1=highest income quintile, 5=lowest income quintile; (c) <u>Household wealth</u>: 1=assets in a home/land, in a car, and in financial investments, 2=assets in home/land and one other asset (cars or financial investments), 3=assets in home/land only or in two other assets (car and financial investments), 4=assets in a car or in financial investments but not in home/land, 5=no assets; (d) <u>Occupational disadvantage</u>: 1=lowest quintile of score, 5=highest quintile of score; (e) <u>Neighborhood socioeconomic disadvantage</u>: 1=lowest quintile of score, 5=highest quintile of score; (f) <u>Neighborhood social disadvantage</u>: 1=lowest quintile of score, 5=highest quintile of score. We then took the sum of all six elements and divided by the number of non-missing elements (all respondents had at least three elements). Higher index values reflect more disadvantage. As a robustness check, we also created a principal components-based score using factor analysis.

### Lifestyle and health behaviors

Smoking was categorized as never smoker, former smoker, or current smoker. In HRS, smoking status is based on the Center for Disease Control (CDC) classification, which defines a smoker as someone who reports smoking 100 cigarettes or more in their lifetime. Individuals were classified as former smokers in the HRS if they report ever being a smoker but do not currently smoke. In MESA, former smoking status was assigned if individuals quit smoking more than one year ago. Adiposity was assessed using the following CDC classifications of body mass index (weight(kg)/height(m^2^)): underweight or normal weight (BMI≤25), overweight (25<BMI<30), or obese (BMI≥30). Alcohol use was assessed using self-reports of alcohol drinking frequency: non-drinker, ≤2 drinks/day, >2 drinks per day. In HRS, frequency of drinking questions was asked in the context of the past three months if an individual reports consuming alcoholic beverages. In MESA, individuals were asked if they presently drink alcohol, and if so, how many glasses of wine, beer, or liquor/mixed drinks they consume per week. In both MESA and the HRS, we adjusted for participant lifestyle/health behaviors reported at the time of blood collection when DNAm was profiled.

### Parental education

The HRS asks respondents the highest grade of schooling completed by their mother or father in years of school, with possible answers ranging from 0 to 17. For 206 individuals with missing data on mother’s education, and 346 individuals with missing data on father’s education, we used publicly available, peer-reviewed imputed measures of mother’s and father’s education.^97^ Imputations were not available for the most recent cohort of HRS participants who were first interviewed in 2010. For respondents with missing data from this cohort, we used dichotomous controls for missing values in our regression specification to avoid any attrition bias from listwise deletion (mother’s education: n=114; father’s education: n=247).

### Polygenic scores (PGS)

We calculated linear PGS for two epigenetic clock phenotypes: Horvath- EAA (i.e., intrinsic epigenetic age acceleration (IEAA)) and Hannum-EAA (extrinsic epigenetic age acceleration (EEAA)) for European ancestry respondents in MESA and the HRS.^66^ The PGS were constructed by taking a weighted sum across the number of SNPs of the number of reference alleles (zero, one, or two) at that SNP multiplied by the effect size for that SNP. SNP effect sizes were obtained from a recent genome-wide association study (GWAS) of 13,493 individuals.^66^ GWAS summary statistics were clumped for linkage disequilibrium (LD) using PLINK (R^2^=0.1, range=250kb).^98, 99^ We computed PGS using PRSice-2^100^ with a fine-tuned p-value cutoff estimated by PUMAS.^61^ PUMAS utilizes GWAS summary statistics to select the optimal p-value threshold as opposed to existing PGS-tuning methods which require a separate validation dataset that is independent from training/testing samples. Summary statistics-based, 4-fold cross validation with replacement was implemented in PUMAS to select the optimal p-value cutoff that gives the highest predictive !^!^ for each trait. After LD clumping and *p*-value thresholding informed by PUMAS, 18 and 36 SNPs were used to construct the IEAA and EEAA PGS, respectively. In MESA, 31 SNPs were used to construct the EEAA PGS. PGS constructed using the PUMAS threshold outperform (or are not significantly different from) PGS constructed with genome-wide significant SNPs (*p*<5E-08) and PGS constructed with all available SNPs (*p*<1) (**Tables S16 and S17**). PGS were standardized to a mean of zero and a standard deviation of one for regression analyses.

### Genetic principal components

In HRS, we performed principal component analysis (PCA) to infer genetic ancestry of 2,265 unrelated individuals of European descent in the HRS DNAm data. We pruned the genotypes for LD and removed long-range LD regions.^101^ The LD-pruned (R^2^ <0.05) analytic HRS dataset consisted of 61,032 genotyped SNPs. We calculated genetic PCs using flashPCA2.^102^ In MESA, EIGENSTRAT was used to calculate PCs using 718,707 SNPs with MAF>0.05 after removing long-range LD regions.^88^

### Statistical analysis: SES index and epigenetic clock associations

Associations between the eight epigenetic clocks and the SES index were assessed using linear regression models and performed in MESA and HRS using SAS and STATA software, respectively.^103, 104^ Primary analyses assessed the relationship between each epigenetic clock and the SES index. To adjust for multiple-hypothesis testing, we used a Bonferroni-adjusted significance threshold for 16 independent tests (eight tests in two samples) and a family-wise error rate of 0.05 (Bonferroni corrected *p*-value=0.003). If the relationship between a clock and the SES index was significant at the Bonferroni-adjusted p-value, we examined associations between the clock and individual components of the SES index in follow-up analyses. Model one regressions controlled for sex, race/ethnicity, age, and marital status. Analyses in MESA also included random effects for methylation chip and position and adjustments for residual sample contamination with non-monocytes (enrichment scores for neutrophils, B cells, T cells, and nature killer cells). Model two regressions further adjusted for lifestyle/health behaviors (smoking, drinking, and BMI) as potential mediators. To avoid losing observations with missing information on a covariate, we set missing values equal to zero and included an additional dichotomous variable set equal to one if an observation was missing.

### Sensitivity and polygenic score interaction analyses

Sensitivity analyses were performed to confirm significant associations by stratifying the analyses by sex, ancestry, and age group (younger than 65 years old or 65 plus). To account for potential differences across MESA exam sites, additional analyses in MESA adjusted for exam site. In HRS, because DNAm was profiled in whole blood, additional analyses adjusted for the percentage of white blood cells present to ensure results were not driven by cell-type composition (i.e., percent neutrophils, monocytes, lymphocytes, eosinophils, and basophils).

Moderation/interaction analyses with PGSs was conducted separately in participants of European ancestry. Regressions were adjusted for the first 10 PCs of the genetic data matrix to account for population stratification^88^ in addition to demographic and lifestyle behaviors (i.e., model two covariates).

## Data Availability

Access to MESA data and HRS restricted data require approval by MESA/HRS and the IRB. HRS epigenetic clocks and other public use data are available on the HRS website.

## Acknowledgments

MESA and the MESA SHARe project are conducted and supported by the National Heart, Lung, and Blood Institute (NHLBI) in collaboration with MESA investigators. Support for MESA is provided by contracts HHSN268201500003I, N01-HC-95159, N01-HC-95160, N01-HC-95161, N01-HC-95162, N01-HC-95163, N01-HC-95164, N01-HC-95165, N01-HC-95166, N01-HC- 95167, N01-HC-95168, N01-HC-95169, UL1-TR-000040, UL1-TR-001079, UL1-TR-001420, UL1-TR-001881, and DK063491. Funding for SHARe genotyping was provided by NHLBI Contract N02-HL-64278. Genotyping was performed at Affymetrix (Santa Clara, California, USA) and the Broad Institute of Harvard and MIT (Boston, Massachusetts, USA) using the Affymetrix Genome-Wide Human SNP Array 6.0. The MESA Epigenomics & Transcriptomics Studies were funded by NIH grants R01HL101250, R01HL119962, R01DK101921, R01HL135009, and 1RF1AG054474. Funding support for the Neighborhood scales dataset was provided by grant HL071759-01.

The HRS is sponsored by the National Institute on Aging (NIA U01AG009740) and conducted by the University of Michigan. This analysis uses data or information from the Contextual Data Resource (CDR): US Decennial Census and American Community Survey Data, 1990-2018, Version 2.0 as of September 2020, developed by Jennifer Ailshire, Sarah Mawhirter, and Eun Young Choi at the USC/UCLA Center on Biodemography and Population Health, with funding from the National Institute on Aging (R21 AG045625, P30 AG017625).

